# Effects of ignoring anaerobic contributions when measuring change in the energy cost of walking in chronic stroke

**DOI:** 10.1101/2025.08.13.25333627

**Authors:** Christina Garrity, Darcy S. Reisman, Sandra A. Billinger, Daniel Carl, Pierce Boyne

**Affiliations:** Department of Rehabilitation, Exercise and Nutrition Sciences, College of Allied Health Sciences, University of Cincinnati, Cincinnati, OH; Department of Physical Therapy, University of Delaware, 540 S, College Ave, Newark, DE; Department of Neurology, School of Medicine, University of Kansas Medical Center, Kansas City, KS; Department of Cell Biology and Integrative Physiology, School of Medicine, University of Kansas Medical Center, Kansas City, KS; University of Kansas Alzheimer’s Research Disease Center, Fairway, KS; Department of Physical Medicine and Rehabilitation, School of Medicine, University of Kansas Medical Center, Kansas City, KS

**Author notes:** **Corresponding author:** Christina Garrity, PT, DPT, NCS. ClinicalTrials.gov Identifier: <u>NCT03760016</u>.

**Keywords:** Gait, Stroke, Oxygen Consumption

## Abstract

**Objective:** To determine whether the common practice of estimating energy cost of walking (CW) using only aerobic energy (oxygen consumption, O_2_) while excluding anaerobic energy (excess carbon dioxide production, CO_2_) introduces systematic and/or random error into the CW change estimates in individuals with chronic stroke.

**Design, setting, and participants:** The HIT-Stroke Trial randomized 55 individuals with walking limitations in chronic stroke to moderate or high intensity walking training.

**Intervention:** Both groups performed walking exercise 3 times per week for 12 weeks.

**Main Outcome:** Treadmill graded exercise testing with metabolic data collection was performed after 0, 4, 8, and 12 weeks of walking training. Mean CW changes (averaged across 4, 8 and 12 weeks) were estimated using only O_2_ (CW_O2_) versus using both O_2_ and CO_2_ (CW_O2+CO2_). CW changes were calculated during the final 3 minutes of each exercise test (i.e. *peak-speed*) and during equivalent speeds of the last 3 minutes of the shortest exercise test (i.e, *matched-speed*). Linear mixed-effects models and bootstrapping were used to compare CW change estimates and their standard errors (SE) between CW_O2_ and CW_O2+CO2_.

**Results:** At *peak-speed*, CW_O2_ showed similar changes to CW_O2+CO2_ (difference: −0.04 J/kg/m, 95% CI: −0.09, 0.03), and had the same relative SE (coefficient of variation difference: 0.0% [−0.9, 0.7]). At *matched-speeds*, CW_O2_ underestimated improvements by 18.2% (0.08 J/kg/m, 95% CI: 0.04, 0.15) compared to CW_O2+CO2_, and had higher relative SE (+10.3% [5.7, 13.5]).

**Conclusion:** Disregarding anaerobic contributions (excess CO_2_) during CW calculation may result in underestimation of training-related improvement and reduce measurement precision when estimating CW changes at *matched-speeds*.

## INTRODUCTION

Stroke can cause neuromuscular impairments (e.g. muscle weakness, spasticity, lack of coordination, co-contraction) that result in biomechanical inefficiencies and increase energy demand up to 3 times that of individuals without disability.^1–4^ Additionally, persons with stroke often have decreased aerobic capacity due to chronic deconditioning.^5^ This combination of increased energy demand and reduced capacity to generate energy is associated with decreased walking capacity and limited participation in everyday walking activity.^6,7^ Stroke rehabilitation interventions often aim to reduce this high energy cost of walking (CW), thus it is important to have accurate and precise measurement of CW changes for evaluating intervention effectiveness.

CW is commonly estimated from oxygen consumption (O_2_) during walking. This accounts for energy contributions from aerobic metabolism, but disregards contributions from anaerobic metabolism.^8^ Anaerobic contributions to energy expenditure are greater when energy demands increase towards an individual’s aerobic capacity.^9^ Since stroke is associated with both increased energy demand for walking and reduced aerobic capacity,^1–5^ it is likely that anaerobic contributions to CW are greater in this population. Indeed, the average stroke survivor appears to reach the ventilatory anaerobic threshold even during preferred-speed walking,^1,10,11^ suggesting that many individuals with stroke have substantial anaerobic contributions to CW. Therefore, accurately assessing CW after stroke likely requires a methodology that accounts for both aerobic and anaerobic energy contributions.

To account for these anaerobic contributions, CW estimation can factor in carbon dioxide production (CO_2_), since anaerobic metabolism leads to a disproportionate increase in CO_2_ expiration relative to the O_2_ consumption rate (i.e. excess CO_2_). This primarily occurs because: (1) anaerobic metabolism leads to metabolic acidosis, which releases CO_2_ from blood bicarbonate as part of the buffering process; and (2) metabolic acidosis, circulating CO_2_ and other factors stimulate hyperventilation, which increases CO_2_ diffusion from the blood into the lungs.^12^ It should be noted that other non-metabolic factors can also contribute to hyperventilation (e.g. anxiety, stress, etc.) and thus can also increase expired CO_2_.^12–14^ However, excess CO_2_ primarily reflects anaerobic metabolism.^12^ Thus, incorporating both O_2_ and CO_2_ measurements in CW estimation may be important for accurately capturing both aerobic and anaerobic energy demands.

Given that intervention studies assess CW *changes*, it is also important to consider how estimating only the aerobic contributions to CW (i.e. using only O_2_) might affect CW change estimates. Change estimates that ignore anaerobic (i.e. excess CO_2_) contributions to CW would be biased if those anaerobic contributions differ across time points. Evidence suggests that these anaerobic contributions may indeed change with walking exercise. For example, multiple studies have demonstrated that walking exercise improves aerobic capacity in individuals after stroke,^11,15–17^ which should reduce reliance on anaerobic energy production at a given intensity. CW change estimates based only on O_2_ would disregard these plausible reductions in anaerobic contributions, which could underestimate training-induced improvements in CW.

Thus, the common practice of calculating CW solely from aerobic metabolism (O_2_) measurements could introduce bias (i.e. systematic error) and/or greater imprecision (i.e. random error) into CW change estimates. The direction and magnitude of this potential bias and imprecision have not been previously assessed. Therefore, this study aimed to assess bias and precision of CW change estimation when CW was estimated using O_2_ only, versus using both O_2_ and CO_2_, in individuals with stroke related walking limitations. Additionally, we assessed how CW estimates relate to motor function, walking capacity and daily walking activity when using O_2_ alone versus both O_2_ and CO_2_.

## METHODS

This analysis used available data from the 55 participants in the HIT-Stroke Trial.^16^ The protocol for this multi-center trial was approved by the University of Cincinnati institutional review board and participants provided written informed consent. Participants were randomized into either moderate intensity aerobic training (MAT) or high intensity interval training (HIIT) group and underwent up to 36 sessions (3 times per week for 12 weeks) involving overground and treadmill walking exercise for 45 minutes each session. The MAT group performed continuous walking with speed adjusted to maintain a target heart rate of 40-60% heart rate reserve (HRR). The HIIT group performed repeated 30 second bursts at maximal safe walking speeds alternated with 30-60 second passive rest (standing or seated), targeting above 60% HRR. A treadmill exercise test with metabolic testing was performed at baseline, and after 4-weeks, 8-weeks, and 12-weeks of training. Eligibility criteria are presented in Table 1.

**Table 1.**
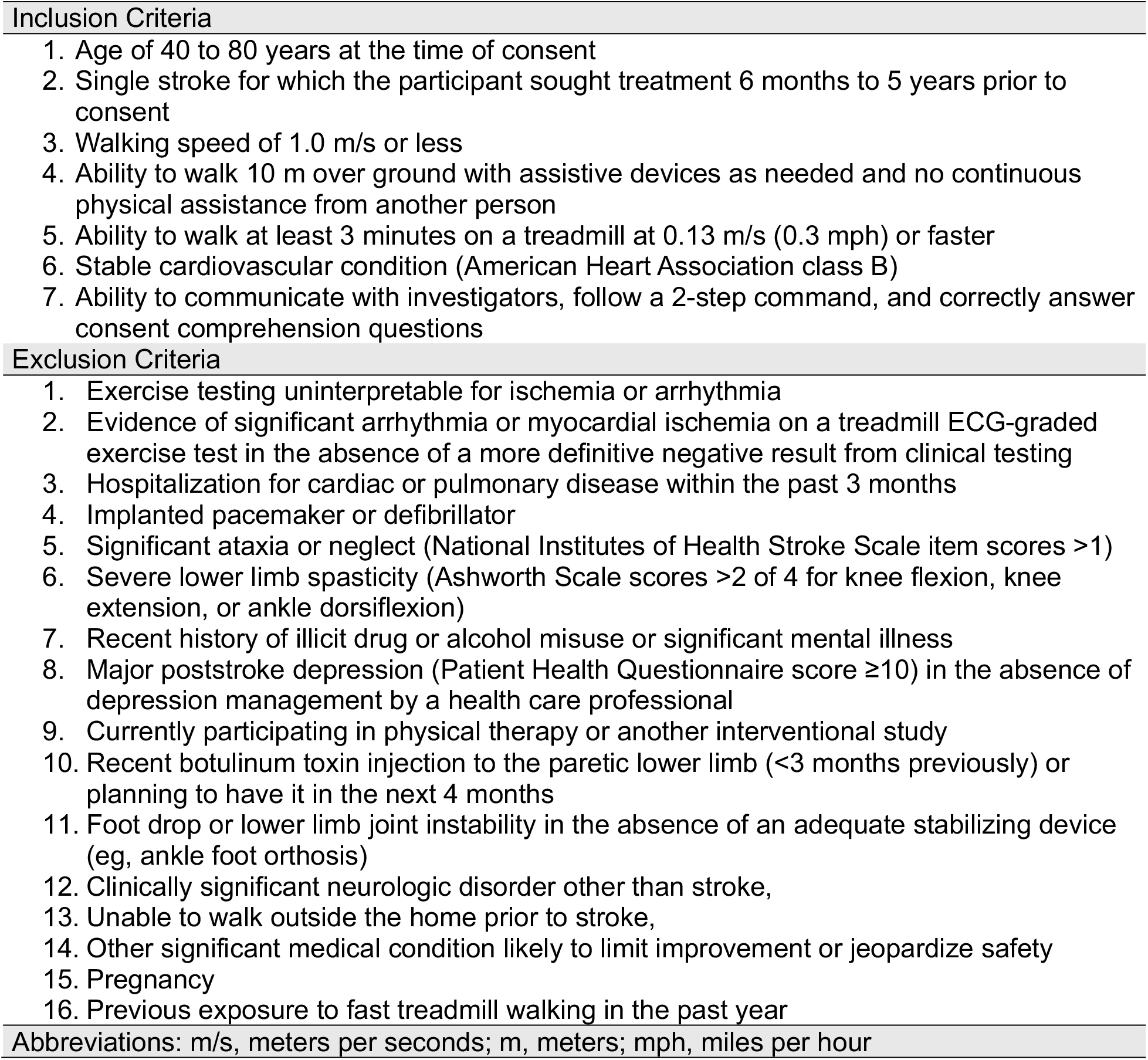
Eligibility criteria.

### Data collection

#### Baseline clinical measures

To examine correlations of baseline CW with motor function, walking capacity, and daily walking activity, several baseline clinical measures were obtained. Motor function was measured using the lower-extremity Fugl-Meyer motor assessment.^18^ Walking capacity was measured using the 6-minute walk test.^19,20^ Baseline daily walking activity was monitored using the Modus StepWatch worn on the nonparetic ankle to determine average steps per day with a goal of recording at least 7 valid days of activity prior to starting the 12 weeks of walking training. Participants were instructed to wear the StepWatch during all waking hours except bathing. An average of 17.5 ± 12.5 days of walking activity were available across participants at baseline.

#### Treadmill exercise testing protocol

Participants walked on the treadmill at 0.3 mph for 3 minutes and then speed increased by 0.1 mph every 30 seconds up to 3.5 mph at which time incline was increased 0.5% every 30 seconds while speed remained fixed at 3.5 mph. Incline data were not used in this analysis, therefore peak workload was limited to 3.5 mph at 0% incline. Test termination occurred if the participant requested to stop, the participant was drifting backward on the treadmill and unable to recover, gait instability was judged to pose an imminent safety risk by the blinded testing therapist, or other stopping criteria according the American college of Sports Medicine guidelines.^21,22^ An overhead safety harness system was used for fall prevention, and participants were able to hold onto the treadmill handrail.

#### Metabolic data collection

To assess walking energetics, O_2_ and CO_2_ data were collected on a breath-by-breath basis (TrueOne 2400, ParvoMedics, Salt Lake City, UT, USA) with a facemask interface during the treadmill exercise test. Participants first performed at least 3 minutes of quiet sitting to achieve a resting steady state. Resting metabolic activity was then averaged across 1 minute of steady state. Due to data collection issues, 3 tests had less than 1-minute averages for resting metabolic activity (40 seconds, 10 seconds, and 22 seconds). Treadmill exercise testing was performed immediately following resting steady state.

### Energy cost of walking calculations

Two different estimates of CW were calculated for comparison,^23^ one that incorporated only O_2_ (CW_O2_) and one that also accounted for both O_2_ and CO_2_ (CW_O2+CO2_).^23^ Seated resting energy expenditure (REST-EE) and walking energy expenditure (WALK-EE) were used to estimate the net energy cost of walking per meter walked for CW_O2_ and CW_O2+CO2_ using the following formulas:

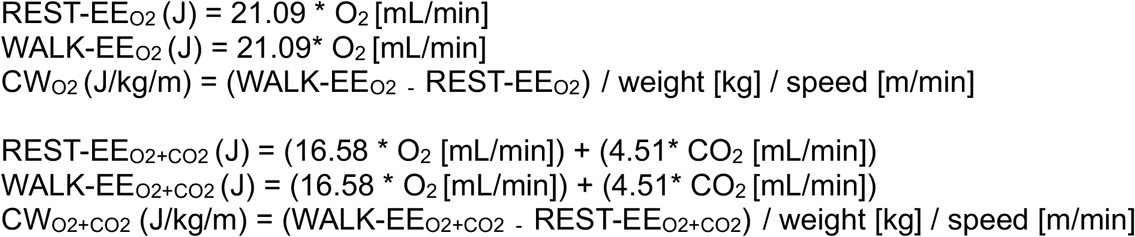

Since CW is known to decrease as walking increases towards non-disabled comfortable speeds,^24^ *change* in CW was evaluated under 2 conditions: *peak-speeds* and *matched-speeds. Peak-speeds* was the average CW across the last 3 minutes of the exercise test (before any incline) from each testing time point (baseline, 4-week, 8-week, and 12-week) allowing CW to be estimated at potentially different speeds across time points. In contrast, the *matched-speeds* condition controlled for speed (i.e. used data collected at the same speeds across testing time points), averaging CW during the last 3 minutes of the shortest exercise test for each participant.

#### Statistical Analysis

To assess how the CW estimates relate to each other and to motor impairment, walking capacity and daily walking activity, Pearson correlations were calculated between baseline CW_O2_, CW_O2+CO2_, lower extremity Fugl-Meyer, 6-minute walk test, and steps per day. Baseline CW was determined using the last 3 minutes of the baseline treadmill exercise test.

Change in CW was estimated using a linear mixed-effects model with fixed effects for time point (baseline, 4-week, 8-week, 12-week), baseline comfortable gait speed (≥0.4 m/s, <0.4 m/s), and study site, with random intercepts and random slopes for each post-baseline time point within participants. This model was selected to resemble the primary statistical model used in HIT-Stroke Trial 1,^16^ except that treatment group was not included because group differences were not relevant to the aims of this analysis. The primary contrast estimated from the model was mean change from baseline across 4-week, 8-week, and 12-week time points.

To assess CW estimation bias, this model estimate for mean change was compared between the CW_O2_ and CW_O2+CO2_ models. To assess differences in precision between the two CW calculation methods, the standard error (SE) for the mean change estimate was compared. To assess differences in the relative precision, the coefficient of variation (CV; SE divided by the mean change estimate) was compared. To obtain 95% confidence intervals for each of these comparisons between models, non-parametric bootstrapping was used, clustered by participant, with the percentile confidence interval method and 5,000 resamples.

The original CW change analysis from this trial only considered aerobic energy contributions and found reduction in peak-speed CW but not matched-speed CW in both groups with no difference between groups.^16^ Therefore, if it was found that disregarding anaerobic contributions introduces bias in CW change estimation, we planned to reanalyze CW change by treatment group accounting for both O_2_ and CO_2_. Based on the results of the analyses above, a sensitivity analysis was then performed to assess whether accounting for anaerobic energy (excess CO_2_) contributions would alter the interpretation of the within-group CW changes or between-group differences previously reported for the HIT-Stroke Trial.

## RESULTS

Baseline participant characteristics are presented in Table 2. Out of the 55 participants enrolled, 10 discontinued participation before the final outcome assessment due to events likely unrelated to the study (including but not limited to COVID-19 shutdown) and 3 participants discontinued due to treatment-related adverse events. One participant required intermittent standing breaks during the treadmill exercise tests for adequate blood pressure safety monitoring therefore continuous walking metabolic data could not be recorded and data could not be used for this analysis.

**Table 2.** Baseline participant characteristics (N=55)

|  |  |
| --- | --- |
| Age, years | 62.4 ± 9.7 |
| Sex, male/female, % male | 36/19 (65.5) |
| BMI | 28.8 ± 4.9 |
| Years since stroke | 2.5 ± 1.3 |
| Use of an assistive device | 34 (61.8) |
| Use of an orthotic | 21.0 (38.2) |
| Functional Ambulation Category >3/5 | 26 (47.3) |
| Fugl-Meyer lower limb motor score, 0-34 | 23.4 ± 5.0 |
| 6-Minute walk test, meters | 239 ± 132 |
| Comfortable gait speed, m/s | 0.64 ± 0.30 |
| Steps per day (n=47) | 4381 ± 2425 |
| Treadmill exercise test duration, minutes | 11.0 ± 5.0 |
| O<sub>2</sub> peak, mL/kg/min | 13.5 ± 4.6 |
| CW<sub>O<sub>2</sub>+CO<sub>2</sub></sub>, J/kg/m (n=54) | 5.54 ± 3.77 |
| CW<sub>O<sub>2</sub></sub>, J/kg/m (n=54) | 4.47 ± 3.04 |

Values reported as mean ± SD or N (%). Variables with n < 55 had missing data at baseline.
Abbreviations: SD, standard deviation; BMI, body mass index; m/s, meters per second, O<sub>2</sub>, oxygen consumption; CW<sub>O<sub>2</sub>+CO<sub>2</sub></sub>, net energy cost of walking calculated using oxygen consumption and carbon dioxide production; CW<sub>O<sub>2</sub></sub>, net energy cost of walking calculated using oxygen consumption.

Correlations between baseline CW estimates, motor function, walking capacity and daily walking activity measures are presented in Table 2. The two CW measures (CW_O2+CO2_ and CW_O2_) were strongly correlated with each other at baseline (r = 1.00) and correlated similarly with lower extremity Fugl-Meyer scores, 6-minute walk test distance, and steps per day, with differences in correlation coefficients between the two CW estimates all ≤ 0.01.

At *peak-speeds* (i.e. measured during the fastest possible speed during each testing time point), CW mean change from baseline for CW_O2_ was −1.01 J/kg/m (p < 0.001, SE = 0.25, CV = 24.8%) and for CW_O2+CO2_ was −0.97 J/kg/m (p < 0.001, SE = 0.24, CV = 24.7%). CW_O2_ did not differ from CW_O2+CO2_ estimation with only 4.0% difference in CW estimation (−0.04 J/kg/m, bootstrapped 95% CI: −0.09, 0.03) and similar absolute SE (0.01 J/kg/m, bootstrapped 95% CI: 0.00, 0.02) and relative SE with a CV difference of 0.0% [−0.9, 0.7] (Table 3).

**Table 3.** Baseline correlations between energy cost of walking estimates and clinical measures, n=54.

| Baseline measure | CW<sub>O2</sub> (r) | CW<sub>O2+CO2</sub> (r) |
| --- | --- | --- |
| CW<sub>O2+CO2</sub> | 1.00* | 1.00* |
| CW<sub>O2</sub> | 1.00* | 1.00* |
| Lower Extremity Fugl-Meyer | -0.71* | -0.70* |
| 6-Minute Walk Test | -0.64* | -0.64* |
| Steps per Day (n=46) | -0.45* | -0.44* |

\*p-value ≤0.002,
**Abbreviations:** CW<sub>O2</sub>, net energy cost of walking calculated using oxygen consumption (J/kg/m); CW<sub>O2+CO2</sub>, net energy cost of walking calculated using oxygen consumption and carbon dioxide production (J/kg/m).

At *matched-speeds* (i.e. measured at the same speed across testing time points), CW mean change from baseline for CW_O2_ was −0.44 J/kg/min (p = 0.14, SE = 0.24, CV = 54.5%) and for CW_O2+CO2_ was −0.52 J/kg/min (p = 0.06, SE = 0.23, CV = 44.2%). Compared with CW_O2+CO2_, CW_O2_ underestimated the CW reduction by 18.2% (0.08 J/kg/m, bootstrapped 95% CI: 0.04, 0.15), had a lower absolute SE by 0.01 J/kg/m, [−0.00, 0.01], and had a higher relative SE with a CV difference of 10.3% [5.1, 13.5] (Table 4).

**Table 4.** Differences in estimated *change* in energy cost of walking (CW) when calculating CW using oxygen consumption alone versus both oxygen consumption and carbon dioxide production. Change averaged across 4-weeks, 8-weeks, and 12-weeks of gait training.

|  | Peak-speed CW<br>(J/kg/m) | Matched-speed CW<br>(J/kg/m) |
| --- | --- | --- |
| $\Delta CW_{O2}$ | -1.01 ± 0.25 (24.8%)<br>[-1.59, -0.43] | -0.44 ± 0.24 (54.5%)<br>[-0.99, 0.11] |
| $\Delta CW_{O2+CO2}$ | -0.97 ± 0.24 (24.7%)<br>[-1.53, -0.41] | -0.52 ± 0.23 (44.2%)<br>[-1.06, 0.01] |
| **Contrasts: $\Delta CW_{O2}$ - $\Delta CW_{O2+CO2}$ differences** |  |  |
| **Bias: Difference in estimate** | -0.04 [-0.09, 0.03]<sup>†</sup> | 0.08 [0.04, 0.15]<sup>†</sup> |
| **Absolute precision: Difference in SE** | 0.01 [0.00, 0.02]<sup>†</sup> | 0.01 [-0.00, 0.01]<sup>†</sup> |
| **Relative precision: Difference in coefficient of variation (SE/estimate)** | 0.0% [-0.9, 0.7] | 10.3% [5.1, 13.5] |

Values are estimates ± SE (% coefficient of variation) [95% CI]
<sup>†</sup>95% CI from bootstrapping
Abbreviations: CW, net energy cost of walking; $\Delta$ , change; CW<sub>O2</sub>, net energy cost of walking calculated using oxygen consumption; CW<sub>O2+CO2</sub>, net energy cost of walking calculated using oxygen consumption and carbon dioxide production; CW<sub>CO2</sub>, net energy cost of walking calculated using only carbon dioxide production; CI, confidence interval; SE, standard error.

Estimating CW change by treatment group revealed that CW_O2_ consistently underestimated improvement in CW following training in both the HIIT and MAT groups at *matched*-*speeds* but not *peak-speeds* (Table 5). Compared with the CW change estimation that only considered aerobic contributions (CW_O2_), factoring in the anaerobic contributions (CW_O2+CO2_) changed the interpretation of within-group differences, but not the between group differences. Within the moderate intensity group, CW_O2+CO2_ showed statistically significant changes not shown by CW_O2_, with 19.6% more improvement at *matched-speeds*.

**Table 5.**
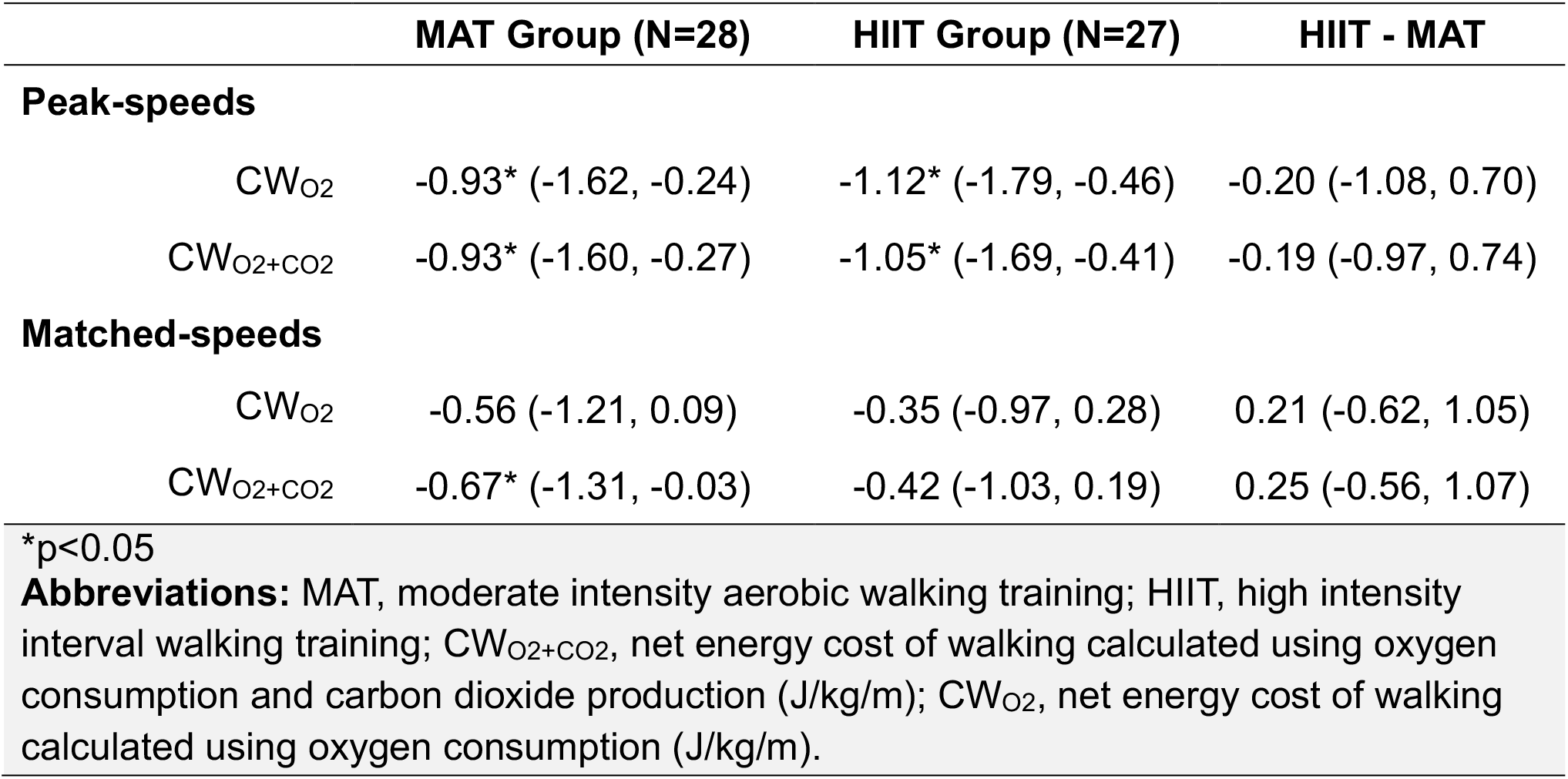
Sensitivity Analysis of HIT-Stroke Trial results by group based on CW calculation method. Values are model estimates [95% CI] of mean change from baseline across 4-week, 8-week, and 12-week time points, n=55

|  | **MAT Group (N=28)** | **HIIT Group (N=27)** | **HIIT - MAT** |
| --- | --- | --- | --- |
| **Peak-speeds** |  |  |  |
| CW<sub>O2</sub> | -0.93* (-1.62, -0.24) | -1.12* (-1.79, -0.46) | -0.20 (-1.08, 0.70) |
| CW<sub>O2+CO2</sub> | -0.93* (-1.60, -0.27) | -1.05* (-1.69, -0.41) | -0.19 (-0.97, 0.74) |
| **Matched-speeds** |  |  |  |
| CW<sub>O2</sub> | -0.56 (-1.21, 0.09) | -0.35 (-0.97, 0.28) | 0.21 (-0.62, 1.05) |
| CW<sub>O2+CO2</sub> | -0.67* (-1.31, -0.03) | -0.42 (-1.03, 0.19) | 0.25 (-0.56, 1.07) |

\*p<0.05
**Abbreviations:** MAT, moderate intensity aerobic walking training; HIIT, high intensity interval walking training; CW<sub>O2+CO2</sub>, net energy cost of walking calculated using oxygen consumption and carbon dioxide production (J/kg/m); CW<sub>O2</sub>, net energy cost of walking calculated using oxygen consumption (J/kg/m).

## DISCUSSION

This study aimed to assess the bias and precision of estimating CW changes based only on aerobic metabolism (i.e. CW_O2_) versus aerobic and anaerobic metabolism (i.e. CW_O2+CO2_) in individuals with stroke related walking limitations. The two methods showed nearly identical correlations with measures of motor impairment, walking capacity and daily walking activity, suggesting that cross-sectional associations are not meaningfully affected by ignoring anaerobic contributions to CW. Estimating CW change at *matched-speeds* based on aerobic metabolism alone did not introduce significant bias and had similar precision to CW change estimation that accounted for both aerobic and anaerobic metabolism. However, when estimating CW change at *matched-speed* based on energy contributions from only aerobic metabolism resulted in significant bias that underestimated the CW reduction. Additionally, CW estimation using aerobic metabolism alone resulted in less relative precision at *matched-speeds* indicating greater measurement error when anaerobic energy demands are not considered. These results indicate that accounting for anaerobic metabolism (i.e. excess CO_2_) when estimating CW changes at *matched speeds* may reduce bias and increase precision for individuals’ post-stroke.

The finding that CW_O2_ underestimated *matched-speed* CW reductions relative to CW_O2+CO2_ provides evidence that anaerobic contributions to CW decreased after walking training. By only using O_2_, the CW_O2_ equation makes the assumption that anaerobic contributions change proportionally to aerobic contributions, while the CW_O2+CO2_ equation allows aerobic and anaerobic contributions to vary independently. Thus, at *matched-speeds* CW_O2_ appears to underestimate CW because it does not capture this disproportionate reduction anaerobic energy contributions. Anaerobic contributions may have decreased because walking training improved the ventilatory threshold and/or overall walking effciency.^16^ Interestingly, this change was only evident with *matched-speed* testing and not *peak-speed* testing. We suspect that this may be because *matched-speed* CW values are taken at a more submaximal workload after training, thus lowering the relative anaerobic demand (excess CO_2_) compared to baseline. In contrast, *peak-speed* CW values are always taken at peak workload, which likely generates similar relative aerobic and anaerobic demands at all time points.

Disregarding anaerobic energy when estimating CW change from baseline may also reduce measurement precision in some circumstances. Compared with CW_O2+CO2_, CW_O2_ had lower relative precision (10.3% higher CV) for assessing CW changes at *matched-speeds*, but not at *peak-speeds*. While absolute precision was no different (similar SE) at *matched-speeds*, this appeared to be related to a lower magnitude of CW change estimates. Since test statistics are based on the magnitude of an estimate *relative* to its SE,^25^ the *relative* SE (CV) is more relevant to statistical power compared with absolute SE. Hence, the lower relative precision of CW_O2_ for estimating *matched-speed* changes could reduce statistical power for these estimates.

CW changes by treatment group indicated that accounting for anaerobic CW contributions (excess CO_2_) was impactful enough to alter the interpretation from the original analysis, which was based solely on aerobic CW (CW_O2_) contributions. While there remained no significant between-group differences in CW change when accounting for anaerobic CW contributions, there was a newly identified significant CW improvement within the MAT group. This is the first CW analysis performed on a longitudinal walking trial using CW methodology that accounts for both aerobic and anaerobic energy contributions, and the first to find significant improvement in *matched-speed* CW in individuals’ post-stroke. Thus, it is possible other previous stroke studies may have also underestimated CW changes following training.

### Study Limitations

Inferring CW based on metabolic biproducts is the most common method for determining CW. However, as an indirect measure, it has limitations. While anaerobic metabolism leads to excess CO_2_ expiration via metabolic acidosis and hyperventilation, non-metabolic factors can also influence hyperventilation (e.g. anxiety, stress, etc.) and thus expired CO_2_.^13,14^ Hyperventilation during exercise can also be influenced by baseline CO_2_ blood concentrations, but this has less influence on CO_2_ measures during slower incremental ramp up exercise testing such as the one used in this trial.^16^ Lastly, individuals with stroke commonly require handrail use during treadmill walking which reduces the metabolic energy cost of walking.^26^ To control for this, hand-support was standardized and similar across testing time points.^26^ Therefore we do not believe this influenced interpretation of the results.

## CONCLUSIONS

CW change estimation using only aerobic energy contributions appears to underestimate CW changes at *matched-speeds*, especially in individuals with greater CW improvement. Relative precision of CW change also decreases when anaerobic contributions are disregarded. Future studies assessing change in CW should consider both aerobic and anaerobic energy contributions to improve accuracy and precision of CW estimates after stroke.

## Data Availability

NICHD Data and Specimen Hub (DASH) DOI: 10.57982/06fb-wh62

https://dash.nichd.nih.gov/study/424597

